# Nutritional traits in the prodromal phase of Parkinson’s disease in community-dwelling older adults in Japan

**DOI:** 10.1101/2023.08.24.23294585

**Authors:** Takashi Yamaguchi, Ryota Nozaki, Keita Taguchi, Yoshio Suzuki, Kai Takahashi, Kenta Takahashi, Kazuhiro Iwaoka, Yuriko Sato, Takahiro Terauchi, Hiroshi Akasaka, Naoki Ishizuka, Kazuhiro Uchida, Toshiharu Ninomiya, Tetsuya Maeda

**Affiliations:** Division of Neurology and Gerontology, Department of Internal Medicine, School of Medicine, Iwate Medical University, 1-1-1 Idaidori, Yahaba, Shiwa, Iwate, Japan 0283694; Department of Health Promotion, School of Health and Nutrition Sciences, Nakamura-Gakuen University, 5-7-1 Befu, Jonan-ku, Fukuoka, Japan 8140198; Department of Epidemiology and Public Health, Graduate School of Medical Sciences, Kyushu University, 3-1-1 Maidashi, Higashi-ku, Fukuoka, Japan 8128582

## Abstract

The growing aging population means that increasing numbers of people are living with Parkinson’s disease (PD). Although dopamine replacement is effective, there are currently no curative or disease-modifying treatments. Nutrition is a well-known environmental risk factor of PD. This study aimed to clarify the relationship between nutritional traits and the prodromal phase of PD.

Subjects were enrolled from community-dwelling older adults. To identify individuals with prodromal PD (PPD), probability of PPD (PPP) and mild parkinsonian sign (MPS) assessments were used. Nutritional status was evaluated using a self-administered food frequency questionnaire form. Intakes of total energy, the three major nutrients (fat, protein, and carbohydrate), B vitamins, water, and dietary fiber were calculated. Nutrient intake in PPD was analyzed using analysis of covariance with age and sex as covariates.

We enrolled 305 subjects. The prevalence of older adults living with PPD was 4.1% by PPP ≥ 0.3 and 21.0% by MPS criteria. Compared with subjects without PPD, individuals with PPP ≥ 0.3 had significantly higher fat intake (33.4 ± 8.8 vs. 29.3 ± 6.7 g/1000 kcal/day, p=0.037) and lower carbohydrate intake (121.4 ± 19.0 vs. 132.1 ± 17.7 g/1000 kcal/day, p=0.039), and MPS-positive subjects had significantly lower total energy intake (1817.5 ± 384.7 vs. 1933.7 ± 433.7 kcal/day, p=0.012).

In this community-based cross-sectional study, nutritional traits differed between older individuals with and without PPD; these differences were more pronounced when the PPP was used to evaluate PPD. Nutrition and diet may thus modify PD incidence.

## Introduction

Parkinson’s disease (PD) is a common neurodegenerative disease in older adults. In the early clinical phase, it is characterized by cardinal movement features such as bradykinesia, rest tremor, and rigidity. Because of the growing aging population worldwide, the number of people living with PD is increasing every year [1]; this global trend has been described as a “pandemic” [2]. Advances in symptomatic treatments based on dopamine replacement have contributed to reducing the mortality rate in PD; however, no curative or disease-modifying treatments have yet been developed [3,4]. Reducing the risk of PD development and preventing its onset is therefore desirable. Motor symptoms reportedly manifest when around 40% of dopamine neurons in the substantia nigra pars compacta have been lost [5]. When motor symptoms are recognized, PD is thus around the middle of its clinical course. However, PD can also present variable non-motor symptoms such as autonomic dysfunction, mood disorders, olfactory impairment, and cognitive decline. These non-motor symptoms occur prior to movement symptoms, and some are regarded as prodromes of PD, with relatively high levels of evidence. People who are living with prodromes only are defined as having prodromal PD (PPD).

Multiple environmental factors are reportedly associated with PD development. Of these, nutrition and diet are well known [6]. Moreover, nutritional habits over the years are modifiable risk factors for many chronic diseases that deteriorate quality of life in both young and older adults. Although many food-derived nutrients and variable dietary patterns are associated with PD development and modification [7], the characteristics of nutrition and diet have not yet been adequately studied in PPD. It has been reported that nutritional characteristics of the Mediterranean diet may be associated with PPD and PD incidence as well as the probability of PPD (PPP) in older adults in the Mediterranean [8]; a study of the nutritional and dietary characteristics of community-dwelling older adults may thus help to elucidate the nutritional risk factors associated with developing PD. Important findings have also been accumulated from the aspect of nutritional absorption. Vagotomy [9] is reportedly a negative risk factor for PD, whereas inflammatory bowel disease [10] is a positive risk factor. Additionally, the specific characteristics of gut microbiota in PD [11] and PD with motor complications [12] are now clear. Nutrient absorption is therefore expected to continue to attract interest in relation to nutrition and diet research.

To detect PPD for research purposes, the PPD research criteria (PPC) was established by the International Parkinson and Movement Disorder Society (MDS) [13]. These criteria encompass various markers that are recognized risk factors of PD. Mild parkinsonian sign (MPS) is another prodromal criteria for detecting subtle parkinsonism that does not meet the diagnostic criteria for PD. MPS has also been used to detect PPD [14,15], and is associated not only with PD development, but also with mortality, aging, dementia, and vascular risk [16].

In the present study, we aimed to clarify the relationship between nutritional status and PPD in community-dwelling older adults.

## Methods

### Subjects

We enrolled subjects from the Yahaba Active Aging and Healthy Brain (YAHABA) study, which has a register of community-dwelling older adults aged 65 years or older. The study details have been described previously [17]. According to the study protocol [18], educational history, past and present history of illness, smoking habits, alcohol consumption, and activities of daily living were comprehensively collected, and physical examinations, neuropsychological and biochemical tests, and radiological examinations were performed in all subjects. Those diagnosed with dementia, PD, or serious systemic diseases were excluded.

### Detection of PPD

We detected PPD using two criteria. First, as in our previous study, PD prodromes were assessed using the PPD self-administered questionnaire (PPQ) and the PPP was calculated [17]. Briefly, the PPQ consists of 21 items that are based on the PPC, including regular pesticide exposure, occupational solvent exposure, non-use of caffeine, current smoking, never smoking, former smoking, first-degree relative with PD, history of type 2 diabetes mellitus, physical inactivity, history of low plasma urate level, daytime somnolence, hyposmia, constipation, urinary dysfunction, impotence for males, neurogenic orthostatic hypotension, symptomatic orthostatic hypotension, depressive mood, and global cognitive deficit. Some items in the PPC are difficult to determine by self-administration; we therefore defined rapid eye movement sleep behavior disorder as the presence of both speech and limb movement during sleep. Ultrasound echogenicity findings of the substantia nigra, dopaminergic positron emission tomography/single-photon emission computed tomography findings, gene testing, and motor markers were excluded. The PPP was calculated using the PPD calculator supplied to MDS members as an online program on the MDS website. We defined PPD in the current study as PPP ≥ 0.3, in accordance with the MDS declaration of research operating policy. The PPQ has been used in a previous retrospective study of pre-onset subjective symptoms in people living with PD, in which prodromes were experienced at an epidemiologically valid frequency [19].

Second, we evaluated MPS [20] as the other criteria for PPD. MPS comprises items of speech, facial expression, rest tremor (rated separately in the lower jaw, right arm, left arm, right leg, and left leg), rigidity (rated separately in the neck, right arm, left arm, right leg, and left leg), standing posture, and general bradykinesia. For MPS, a neurologist performed a neurological examination and rated each item from 0 (none) to 4 (severe). MPS positivity was defined when any one of the following criteria was satisfied: (1) at least two items with scores ≥ 1, (2) at least one item with a score ≥ 2, or (3) at least one item of rest tremor with a score ≥ 1.

### Nutritional status

We assessed nutritional status using a self-administered food frequency questionnaire (FFQ) form according to a previous protocol [18]. This questionnaire is a booklet-type form with illustrations and photographs of portion sizes and is easy for older adults to complete. Support from family members was acceptable, and cohort staff who were independent of the study checked for discrepancies and missing values. The FFQ assesses the consumption frequency of 233 foods, food groups, or beverages during the preceding month, and can evaluate the intake of 23 food groups and 160 nutrients. In the present study, we analyzed the intakes of total energy, the three major nutrients (fat, protein, and carbohydrate), some vitamins (B1, B2, B6, and B12), water derived from food, and dietary fiber. Total energy intake was examined in terms of the bare minimum; other intakes were examined using the residue method.

A validation study of this FFQ has been conducted in 60 residents (aged 65 years or older) of Hisayama, located on Kyushu island in Japan, who participated in an annual health checkup in 2019 and consented to participate in this study. Dietary surveys were performed using a weighing and dietary recording method for 4 consecutive days (3 weekdays + 1 weekend day) for a total of 16 days over four seasons (November–December 2019, February–March 2020, May–June 2020, and August– September 2020). Subjects first completed the FFQ at a health center or at home. They were then provided with the dietary recording form, recording manual, digital weighing scale, measuring spoons, a camera photography scale (a standard graphic tool used in the National Health and Nutrition Survey), and an instant camera (if unable to take pictures on a device such as a mobile or smartphone). Nutritional values were calculated according to the Japanese Standard Tables of Food Composition 2015 (7th revision) [21]. The validity of each food and nutrient intake calculated from the FFQ was compared with that calculated from the dietary surveys (the average intake of each food/nutrient from the 16-day period over four seasons) using Pearson’s correlation coefficients. The results were as follows: 0.66 for total energy, 0.59 for fat, 0.61 for protein, 0.49 for carbohydrate, 0.56 for vitamin B1, 0.59 for vitamin B2, 0.37 for vitamin B6, 0.31 for vitamin B12, 0.47 for water derived from food, and 0.66 for dietary fiber (all p<0.01). Information regarding these data is presented briefly on the JPSC-AD website (https://www.eph.med.kyushu-u.ac.jp/jpsc/link/pdf/ffq02.pdf). In addition, the relevant manuscript of this validation study is currently in preparation.

### Statistical analysis

We calculated the bivariate associations of PPP and MPS with the clinical backgrounds of subjects using the Mann–Whitney U test for nonparametric variables and chi-squared or Fisher’s exact tests for categorical variables. We evaluated the relationship between PPD (i.e., PPP ≥ 0.3 or MPS positivity) and nutrients using an analysis of covariance, with age, sex, and constipation added as covariates. In the current study, sex was taken to mean the sex assigned at birth. P-values < 0.05 were considered significant. All analyses were performed using IBM SPSS Statistics for Windows (version 23.0; IBM Corp., Armonk, NY, USA).

### Ethical approval

All investigators conducted this study in accordance with the Declaration of Helsinki. The research protocol was approved by the Ethics Committee of Iwate Medical University (HGH28-12, HG2020-017) and was performed with written informed consent from all subjects.

## Results

### Subject demographics

We targeted 962 older adults who were registered in the YAHABA study. Responses to the PPQ were obtained from 715 subjects and MPS scores were obtained from 345 subjects. We finally enrolled 305 subjects in the study, for whom both PPQ and MPS data were available. Table 1 shows the background demographics of subjects. The prevalence of older adults living with PPD was 3.9% using PPP ≥ 0.3 and 21.0% using MPS criteria (i.e., prevalence was around five times higher with MPS criteria than with PPP ≥ 0.3). In subjects with PPP ≥ 0.3, Geriatric Depression Scale scores were significantly higher than in subjects with PPP < 0.3 (6.67 ± 4.10 vs. 3.24 ± 2.97, p=0.02). Furthermore, MPS-positive subjects were significantly older (78.7 ± 6.2 vs. 76.2 ± 5.1 years, p=0.004) and had higher Geriatric Depression Scale scores (4.70 ± 3.35 vs. 3.05 ± 2.93, p=0.004) than MPS-negative subjects. Sex; medical histories of stroke, myocardial infarction, and type 2 diabetes mellitus; Mini-Mental State Examination score; body mass index; and walking speed were not significantly associated with PPP or MPS scores. The scores and frequencies for each of the MPS subitems are shown in Table 2. In the community-dwelling older adults enrolled in the present study, posture and rigidity had relatively high scores and frequencies, whereas tremor had relatively low scores and frequencies.

**Table 1.** Subjects’ demographics.

|  | All<br>(N = 305) | Prodromal PD<br>probability |  | Mild parkinsonian sign |  |
| --- | --- | --- | --- | --- | --- |
| | | $\geq 0.3$<br>(N=12) | $< 0.3$<br>(N=293) | Positive<br>(N=64) | Negative<br>(N=241) |
| Sex, male:female, n | 140:165 | 5:7 | 134:159 | 34:30 | 106:105 |
| Age, mean ± SD,<br>years | 76.93 ±<br>5.60 | 76.67 ±<br>6.42 | 76.69 ±<br>5.35 | 78.64 ±<br>6.20* | 76.24 ±<br>5.05 |
| Stroke, n (%) | 25 (8.2) | 2 (16.7) | 23 (7.8) | 8 (12.5) | 17 (7.1) |
| Malignant<br>neoplasm, n (%) | 54 (17.7) | 1 (8.3) | 53 (18.1) | 13 (20.3) | 41 (17.0) |
| Diabetes mellitus, n<br>(%) | 54 (17.7) | 4 (33.3) | 50 (17.1) | 10 (15.6) | 44 (18.3) |
| GDS, mean ± SD | 3.40 ±<br>3.09 | 6.67 ±<br>4.10* | 3.24 ±<br>2.97 | 4.70 ±<br>3.35** | 3.05 ±<br>2.93 |
| MMSE, mean ± SD | 28.13 ±<br>2.47 | 28.42 ±<br>2.50 | 28.10 ±<br>2.56 | 27.73 ±<br>2.65 | 28.24 ±<br>2.41 |
| BMI, mean ± SD | 24.28 ±<br>3.48 | 23.89 ±<br>2.83 | 24.17 ±<br>3.88 | 23.98 ±<br>4.22 | 24.23 ±<br>3.75 |
| Walking speed,<br>mean ± SD, s/5m | 4.56 ±<br>1.10 | 5.15 ±<br>1.27 | 4.54 ±<br>1.09 | 4.87 ±<br>1.57 | 4.48 ±<br>0.94 |

N & n, number; SD, standard deviation; GDS, geriatric depression scale; MMSE, mini mental examination; BMI, body mass index; \*, $p < 0.05$ ; \*\*, $p < 0.01$ .

**Table 2.**
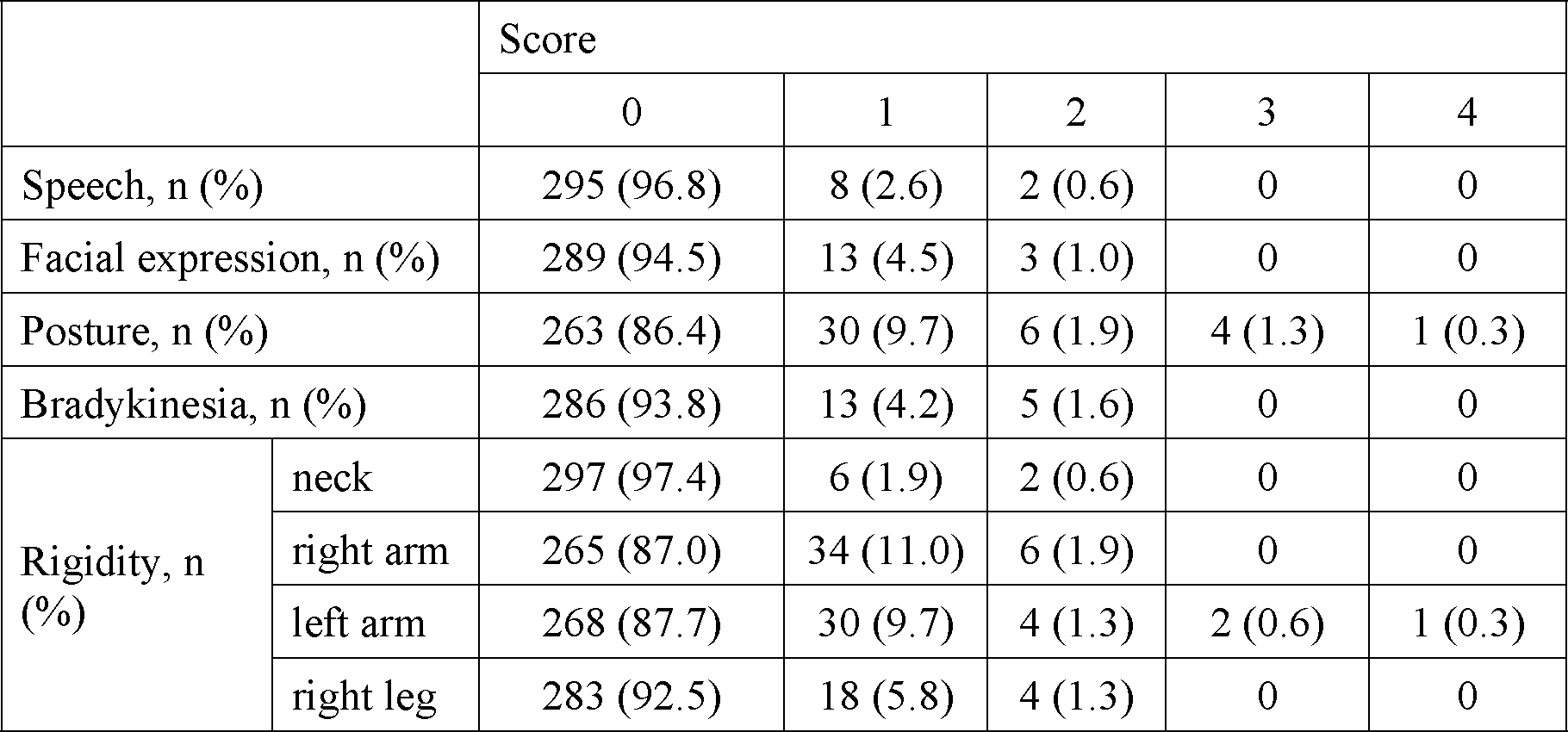
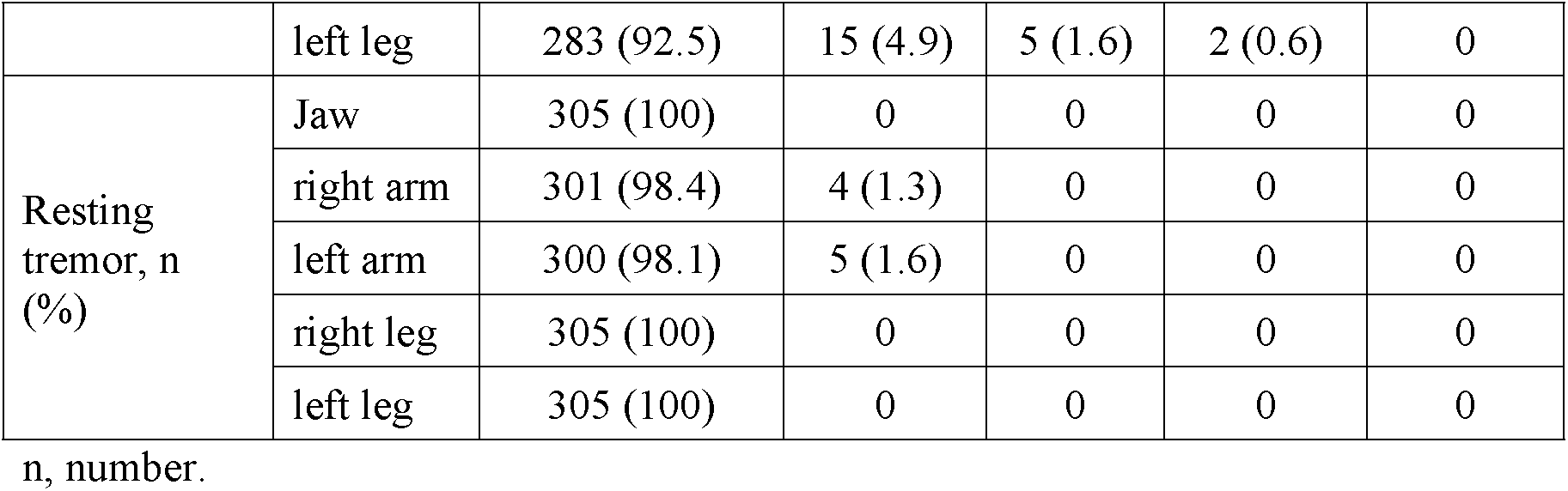
Score of mild parkinsonian sign in this study older population.

|  |  | Score |  |  |  |  |
| --- | --- | --- | --- | --- | --- | --- |
|  |  | 0 | 1 | 2 | 3 | 4 |
| Speech, n (%) |  | 295 (96.8) | 8 (2.6) | 2 (0.6) | 0 | 0 |
| Facial expression, n (%) |  | 289 (94.5) | 13 (4.5) | 3 (1.0) | 0 | 0 |
| Posture, n (%) |  | 263 (86.4) | 30 (9.7) | 6 (1.9) | 4 (1.3) | 1 (0.3) |
| Bradykinesia, n (%) |  | 286 (93.8) | 13 (4.2) | 5 (1.6) | 0 | 0 |
| Rigidity, n<br>(%) | neck | 297 (97.4) | 6 (1.9) | 2 (0.6) | 0 | 0 |
|  | right arm | 265 (87.0) | 34 (11.0) | 6 (1.9) | 0 | 0 |
|  | left arm | 268 (87.7) | 30 (9.7) | 4 (1.3) | 2 (0.6) | 1 (0.3) |
|  | right leg | 283 (92.5) | 18 (5.8) | 4 (1.3) | 0 | 0 |
|  | left leg | 283 (92.5) | 15 (4.9) | 5 (1.6) | 2 (0.6) | 0 |
| Resting tremor, n (%) | Jaw | 305 (100) | 0 | 0 | 0 | 0 |
|  | right arm | 301 (98.4) | 4 (1.3) | 0 | 0 | 0 |
|  | left arm | 300 (98.1) | 5 (1.6) | 0 | 0 | 0 |
|  | right leg | 305 (100) | 0 | 0 | 0 | 0 |
|  | left leg | 305 (100) | 0 | 0 | 0 | 0 |

n, number.

### Differences in nutritional status between PPD defined by PPP and MPS

In Table 3, nutritional status adjusted for age and sex is classified by PPP ≥ 0.3 or < 0.3 and MPS positivity or negativity. Subjects with PPP ≥ 0.3 had no difference in total energy intake compared with those with PPP < 0.3; however, MPS-positive subjects had significantly lower total energy intake than MPS-negative subjects (1817.5 ± 384.7 vs. 1933.7 ± 433.7 kcal/day, p=0.012). Analysis of the three major nutrients showed significantly higher fat intake (33.4 ± 8.8 vs. 29.3 ± 6.7 g/1000 kcal/day, p=0.037) and lower carbohydrate intake (121.4 ± 19.0 vs. 132.1 ± 17.7 g/1000 kcal/day, p=0.039) in subjects with PPP ≥ 0.3 than in those with PPP < 0.3. There were no differences in the intakes of the three major nutrients between subjects with positive and negative MPS. Intake of B vitamins was not different between subjects living with and without PPD, evaluated using either the PPP or MPS criteria. Water intake derived from food tended to be lower in subjects with PPD than in those without PPD, but this result was not significant (771.9 ± 188.9 in subjects with PPP ≥ 0.3 and 844.3 ± 224.5 in subjects with PPP < 0.3, and 803.7 ± 188.5 in MPS-positive subjects and 851.4 ± 254.4 mL/day/1000kcal in MPS-negative subjects). Dietary fiber intake also tended to be lower in subjects with PPP ≥ 0.3 than in those with PPP < 0.3, but this difference was not significant. Constipation had no effect as an additional confounder (data not shown).

**Table 3.** Nutritional traits adjusted for age and sex effects in older adults living with prodromal phase of PD evaluated with PPC or MPS.

|  | Prodromal PD probability |  |  | Mild parkinsonian sign |  |  |
| --- | --- | --- | --- | --- | --- | --- |
| | $\geq 0.3$ | $< 0.3$ | P value | Positive | Negative | P value |
| Total energy intake, kcal/day | 1860.9 ± 312.7 | 1911.5 ± 430.3 | 0.748 | 1817.5 ± 384.7 | 1933.7 ± 433.7 | 0.012 |
| Fat intake, g/1000 kcal/day | 33.4 ± 8.8 | 29.3 ± 6.7 | 0.037 | 29.3 ± 6.0 | 29.9 ± 6.2 | 0.656 |
| Carbohydrate intake, g/1000 kcal/day | 121.4 ± 19.0 | 132.1 ± 17.7 | 0.039 | 134.9 ± 15.2 | 130.8 ± 18.4 | 0.085 |
| Protein intake, g/1000 kcal/day | 39.4 ± 8.8 | 39.3 ± 6.7 | 0.906 | 38.5 ± 5.2 | 39.5 ± 7.1 | 0.263 |
| Vitamin B1 intake, mg/1000 kcal/day | 0.5 ± 0.1 | 0.5 ± 0.1 | 1.000 | 0.5 ± 0.1 | 0.5 ± 0.1 | 0.257 |
| Vitamin B2 intake, mg/1000 kcal/day | 0.7 ± 0.2 | 0.7 ± 0.2 | 0.888 | 0.7 ± 0.1 | 0.7 ± 0.2 | 0.235 |
| Vitamin B6 intake, mg/1000 kcal/day | 2.0 ± 0.9 | 1.9 ± 0.7 | 0.667 | 1.9 ± 0.6 | 1.9 ± 0.7 | 0.535 |
| Vitamin B12 intake, μg/1000 kcal/day | 4.4 ± 2.3 | 4.1 ± 1.6 | 0.539 | 4.0 ± 1.3 | 4.1 ± 1.7 | 0.319 |
| Water intake (food), g/1000 kcal/day | 771.9 ± 188.9 | 844.3 ± 224.5 | 0.290 | 803.7 ± 188.5 | 851.4 ± 254.4 | 0.307 |
| Fiber intake, g/1000 kcal/day | 8.0 ± 2.6 | 8.5 ± 2.4 | 0.398 | 8.4 ± 2.1 | 8.5 ± 2.5 | 0.753 |

PPC, prodromal Parkinson's disease criteria; MPS, mild parkinsonian sign; PD, Parkinson's disease; SD, standard deviation; all values are expressed as mean ± standard deviation.

### Association between prodromal PD criteria and MPS criteria

As shown in Figure 1, there were four subjects with PPP ≥ 0.3 in the MPS-positive group (6.3%) and eight in the MPS-negative group (3.3%). Mean PPP values were 0.066 in the MPS-positive group and 0.047 in the MPS-negative group. There were no significant differences between the two groups in subject number or mean values (p=0.281 and p=0.148, respectively). A comparison of prodrome frequencies between MPS-positive and -negative subjects is shown in Figure 2; the non-use of caffeine was significantly more common in the MPS-positive group than in the MPS-negative group (40.7% vs. 19.4%, p<0.001). However, no other differences were observed.

**Fig 1.**
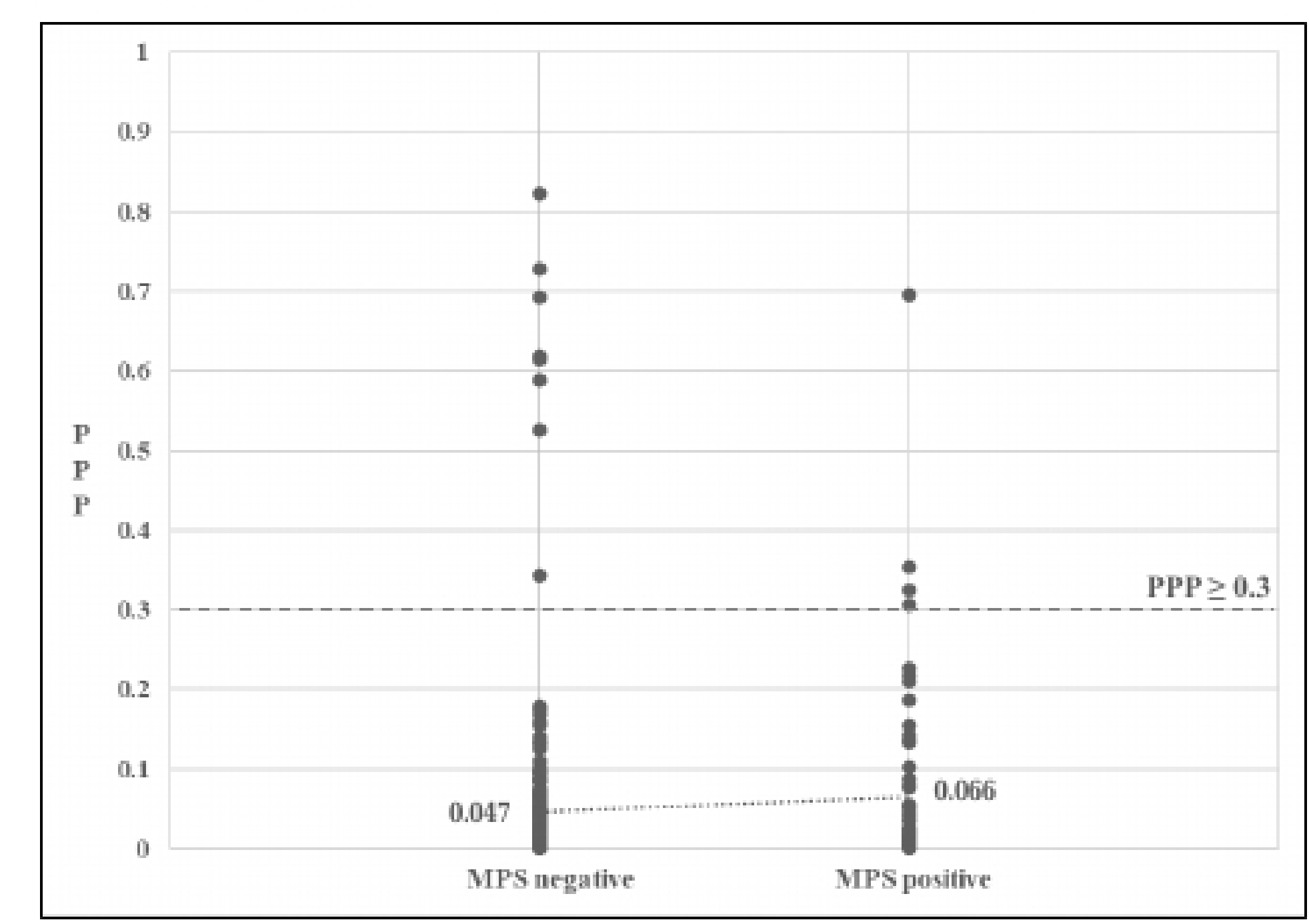
Association between PPP and MPS. The figure shows PPP values when divided between two groups with and without MPS. Mean value of PPP was not significantly different between two groups (0.066 vs 0.047, p=0.140). Dashed line means PPP = 0.3. PPP, probability of prodromal Parkinson’s disease; MPS, mild parkinsonian sign

**Fig 2.**
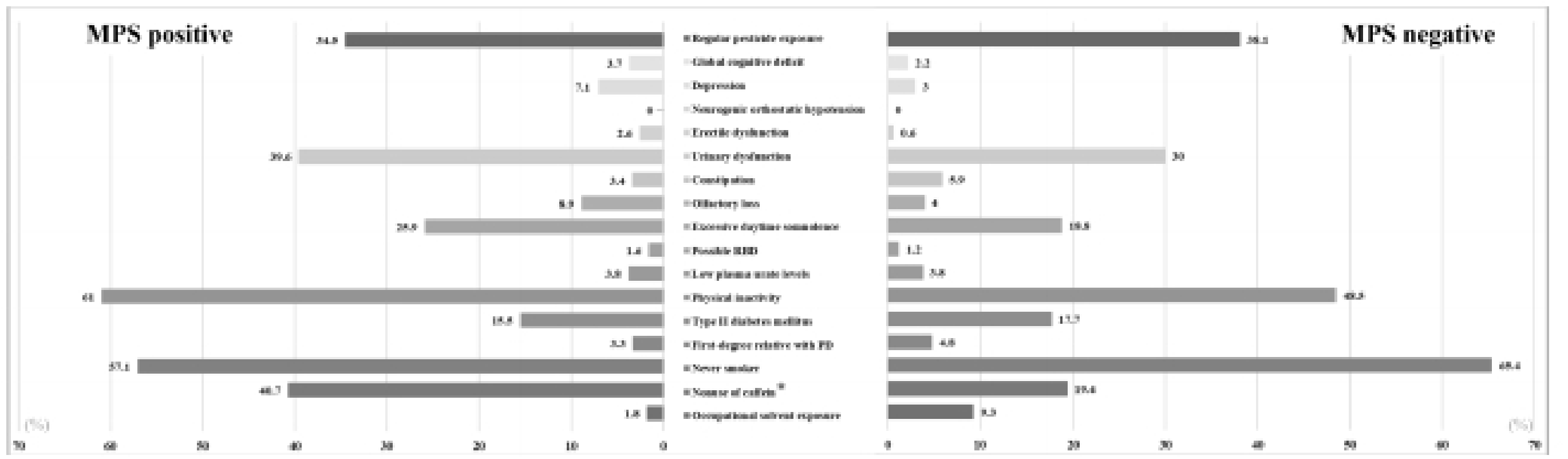
Prodrome features between MPS-positive and -negative older adults. Frequencies of each prodromal item are shown in two groups (MPS positive or MPS negative). The frequency of nonuse of caffeine was significantly higher in the MPS-positive group (p<0.01). MPS, mild parkinsonian sign.

## Discussion

The current community-based cross-sectional study revealed differences in nutritional traits between older individuals with and without PPD. These differences were especially pronounced when older adults were evaluated using the PPQ (based on the PPC). Older adults with PPP ≥ 0.3 exhibited a unique nutritional trait of a higher daily intake of fat and a lower daily intake of carbohydrate compared with those with PPP < 0.3. Moreover, in older adults with positive MPS, a lower total daily energy intake was identified. Because individuals with PPD are a high-risk population for developing PD, the characterization of nutritional traits in PPD in the present study is considered very important. Nutritional intervention in daily diets may contribute to reducing the incidence of PPD, and thus possibly the incidence of PD.

The association between PD development and lipid intake is controversial, as reported in several previous studies [22,23]. Several negative effects of lipid intake have been described in relation to PD pathogenesis, such as effects on α-synuclein aggregation, oxidative stress, and inflammatory cytokines. Moreover, a previous meta-analysis demonstrated that a high intake of animal fat (arachidonic acid and cholesterol) increases PD risk, whereas a high polyunsaturated fatty acid intake decreases the risk [24]. Although elevated cholesterol levels cause oxidative stress and mitochondrial dysfunction [25], positive effects of lipids have also been described. For example, because polyunsaturated fatty acids are present in neuronal cell membranes, fat intake may have a protective effect on neuronal cells. Thus, lower lipid intake may accelerate the development of PD.

There have been few reports of carbohydrate or total energy intake in relation to PD. A meta-analysis reported no independent associations between the dietary intake of carbohydrates or total energy and PD risk [26]. However, it remains unclear whether nutritional status before the beginning of the disease process of PD may be associated with disease development. Nonetheless, the influence of nutritional status on motor function is greater in older adults than in young adults. Thus, because a motor function decline in older adults increases the risk for reduced activities of daily living and quality of life (whether or not PD eventually develops), adequate nutritional management should be recommended, at least from the perspective of general wellbeing. Furthermore, the impact of nutritional status on PD development should be further investigated, and prospective studies should be conducted that integrate nutrients into the research criteria.

Different definitions of MPS have been reported previously [27]. In the present study, we chose to use the criteria reported by Louis et al. [2]] to avoid confusion with the PPC and because we wanted our findings to be comparable with those of a previous study [28]. The MPS positivity rate among community-dwelling older adults was 21.0% in the current study, which was approximately five times higher than the rate of PPD when estimated using PPP ≥ 0.3. Similarly, a previous study from Japan reported a prevalence of MPS positivity of 22.1% in community-dwelling older adults aged 60 years and older [28], and a systematic review has reported prevalence rates of MPS positivity as 4%–46% [16]. The results of the present study are therefore comparable to previous studies and support their results. In our previous investigation [17], the crude prevalence of PPD using PPP ≥ 0.3 was 5034.5/100,000 (5.0%) community-dwelling older adults. Together with the current finding of a PPD prevalence of 3.9% in older adults using PPP ≥ 0.3, it is therefore likely that the MPS criteria detect a broader population of candidates for PPD in community-dwelling older adults than when PPP ≥ 0.3 is used. Indeed, in the present study, the numbers of older adults with PPP ≥ 0.3 or < 0.3 were not different between MPS-positive and -negative older adults. Furthermore, although there were no differences in age between subjects with PPP ≥ 0.3 and < 0.3, MPS-positive subjects were significantly older than MPS-negative subjects. Together, these findings support the idea that MPS criteria are relatively unspecific in older adults [29].

We have previously reported the characteristics of PD prodromes in community-dwelling older adults with PPP ≥ 0.3 [17]. In the current study, however, there were no significant differences in PD prodromes between subjects with PPP ≥ 0.3 and < 0.3. One reason for these different results may be the relatively small number of older adults with PPP ≥ 0.3. By contrast, MPS positivity in older adults was associated with caffeine non-intake. Although caffeine intake is a well-known protective factor for PD onset [30], there is no previous evidence of its association with MPS.

The present study has several limitations. First, it was a single-center study that was limited to a cohort of Japanese subjects. Second, the PPQ needs to be further improved for its future intensive research use. Third, the number of subjects with PPP ≥ 0.3 was relatively small. Finally, the cohort format (where subjects were required to come to the screening site) may have created a selection bias because many of the subjects who underwent screening were likely relatively healthy.

## Conclusions

In the present community-based cross-sectional study, nutritional traits differed between older individuals with and without PPD. When older adults were evaluated using the PPQ (based on the PPC), those with PPP ≥ 0.3 exhibited a higher daily intake of fat and a lower daily intake of carbohydrates than those with PPP < 0.3. The characterization of nutritional traits in older adults is considered very important because nutritional intervention in daily diets may reduce the incidence of PD.

## Data Availability

All data produced in the present study are available upon reasonable request to the authors

## Acknowledgments

We thank Ms. Kanako Abe for general study support. We also thank Bronwen Gardner, PhD, from Edanz (https://jp.edanz.com/ac) for editing a draft of this manuscript.

## Notes

### Competing Interest Statement

The authors have declared no competing interest.

### Funding Statement

This study was supported by Grants-in-Aid from the Japan Agency for Medical Research and Development (JP21dk0207053 and JP21gm1010002).

### Author Declarations

Ethics committee of Iwate Medical University gave ethical approval for this work

